# Distress is positively associated with induced secondary hyperalgesia in people with suppressed HIV

**DOI:** 10.1101/2025.01.27.25321015

**Authors:** Luyanduthando Mqadi, Gillian J. Bedwell, Ncumisa Msolo, Gwendoline Arendse, Maia Lesosky, Peter R. Kamerman, Mark R. Hutchinson, Andrew Schrepf, Robert R. Edwards, John A. Joska, Romy Parker, Victoria J. Madden

**Affiliations:** African Pain Research Initiative, Department of Anaesthesia and Perioperative Medicine,□Neuroscience Institute, University of Cape Town, Cape Town, South Africa□; National Heart and Lung Institute, Imperial College London, London, United Kingdom; Department of Physiology, School of Biomedical Sciences, University of the Witwatersrand, Johannesburg, South Africa; School of Biomedicine, University of Adelaide, South Australia, Australia; Australian Research Council Centre of Excellence for Nanoscale BioPhotonics.; Chronic Pain and Fatigue Research Center, Department of Anesthesiology, Michigan Medicine, University of Michigan, Ann Arbor, Michigan, USA; Department of Anesthesiology, Perioperative, and Pain Medicine, Brigham & Women’s Hospital, Harvard Medical School, Boston, Massachusetts, USA; HIV Mental Health Research Unit, Department of Psychiatry and Mental Health,□Neuroscience Institute, University of Cape Town, Cape Town, South Africa□

**Author notes:** Corresponding author. Postal address: D23 Department of Anaesthesia, Groote Schuur Hospital, Anzio Rd, Observatory, 7925, South Africa. **Previous presentation of this work** Preliminary results: Abstract (2022) and poster (2023) presentations at the PainSA Congress.

**Keywords:** distress, HIV, secondary hyperalgesia, pain-free, persistent pain

## Abstract

Pain and symptoms of depression and anxiety (here, ‘psychological distress’) are frequently reported by people with HIV. Although pain is widely acknowledged to contribute to distress, distress may also contribute to pain and its persistence. Facilitation of nociceptive signalling is one pathway by which distress could exacerbate pain. The current study investigated the relationships between symptoms of depression and anxiety, secondary hyperalgesia (SH), and persistent pain in people with HIV, reporting pain (n=19) or no pain (n=26). We hypothesised that self-reported distress would be positively associated with the surface area (primary measure) and magnitude (secondary measure) of induced SH, and that participants reporting persistent pain would display greater induced SH than those reporting no pain. We found that distress was positively associated with the surface area (p=0.02) and the magnitude (p=0.01) of induced SH. However, participants with persistent pain showed no difference in the surface area of SH compared to pain-free participants (p=0.87), and those with pain displayed a marginally lower magnitude of SH (p=0.05). These findings position SH as a potentially useful mechanistic outcome for interventions that aim to address pain by reducing symptoms of depression and anxiety.

**Perspective:** Symptoms of depression and anxiety were positively associated with induced secondary hyperalgesia in people with suppressed HIV.

## Introduction

Pain is more prevalent in people with HIV than in the general population ^[1–3]^. Similarly, psychological distress—a cluster of negative affective symptoms including symptoms of depression or anxiety ^[4]^—occurs more frequently in people with HIV than in their HIV-negative peers ^[5; 6]^.

There is considerable overlap between pain and psychological distress. Pain-focused concerns, such as diagnostic uncertainty, are linked to distress; people with persistent pain typically score higher on multiple indices of distress ^[4; 7–9]^, and living with pain can exacerbate pre-existing mood problems ^[10; 11]^. In people with HIV, having pain is associated with twice the rate of suicidal ideation ^[12; 13]^. HIV-focused difficulties, such as the shock of an HIV diagnosis, changes in sexual behaviour, lack of support, and the strain of long-term disease management, can also contribute to distress ^[14–17]^.

Psychological distress can also worsen pain and its impacts in people with and without HIV ^[18]^. Although primarily psychological mechanisms are an obvious pathway for this relationship, it is also plausible that distress increases neuronal responsiveness to nociceptive signals in the central nervous system. Given the established links between ‘pain sensitivity’-based tests of neuronal responsiveness and persistent pain, a distress-driven increase in neuronal responsiveness might then contribute to persistent pain ^[19]^.

However, not all people with persistent pain show increased ‘pain sensitivity’ to psychophysical tests; responses vary between individuals and across the different tests, and pooled estimates of the relationship between these tests of pain sensitivity and psychological distress suggest a modest relationship at best ^[19; 20]^. Psychophysical testing in people with HIV and pain has shown increased ‘pain sensitivity’ ^[21; 22]^, possibly arising from sustained exposure to inflammatory stimulants such as the HIV envelope protein gp120 and lipopolysaccharide, even with viral suppression ^[23]^. Such sustained provocation of inflammation can meaningfully alter neuronal responsiveness; for example, by shifting ratios between cytokines, altering neuron-astrocyte interactions, and initiating phenotypic switching of microglia ^[24–26]^. To better understand the mechanistic process that underpins persistent pain in the context of HIV, it is important to clarify the relationships of neuronal responsiveness to psychological distress, and persistent pain, in people living with HIV.

Induced secondary hyperalgesia provides a well characterised model of neuronal responsiveness that is safe to apply, and elicits a short-lived hypersensitivity of the skin that is thought to reflect a spinal heterotopic long-term potentiation-like process, although both facilitatory and inhibitory mechanisms contribute ^[27–29]^. This model has been used to demonstrate a relationship of induced secondary hyperalgesia to negative affect in healthy women with history of stressful life events ^[30]^, with worse surgical outcomes in surgical patients ^[31]^, and with experimentally induced negative affect in healthy participants ^[32]^, but to our knowledge, no study has used this model in people with HIV. Two recent reviews synthesised the small body of evidence and found that psychological manipulations— including placebo/nocebo, attention and cognitive load, social support, cognitive behavioural therapy, threat/fear induction, and emotional disclosure—may modulate secondary hyperalgesia, but none of those studies were done in people with HIV ^[33; 34]^.

This study aimed to clarify the relationship of induced secondary hyperalgesia (SH) to symptoms of depression and anxiety and persistent pain status, in people with HIV. It tested two hypotheses: (i) that symptoms of depression and anxiety are positively associated with induced SH (*primary hypothesis*), and (ii) that people with HIV reporting persistent pain display greater induced SH than people with HIV reporting no pain (*secondary hypothesis*).

## Methods

### Overview

The cross-sectional data used here were collected as part of a larger study focused on psychological distress, inflammatory reactivity, induced SH, and persistent pain in people with HIV (protocol published at ^[35]^). In brief, participants with well controlled HIV, who reported either no pain or persistent pain, who had completed the distress self-report questionnaire, and who were eligible to undergo electrical stimulation, were invited to attend a separate session where SH was induced and assessed.

Ethical approval was granted by the Faculty of Health Sciences Human Research Ethics Committee of the University of Cape Town (764/2019) and the local health authority (ref: 24699). The larger study was registered at ClinicalTrials.gov (NCT04757987). The analysis plans for the current study were posted and locked online along with a preliminary version of the analysis script developed using pilot data, at Open Science Framework (posted on 30 March 2022). An updated analysis plan was locked on 7 April 2022 before data were processed, and a record of blinded analysis and interpretation was locked on 16 November 2022, before unblinding of the analyst (LM). [the link to online project: https://osf.io/2hdpy/?view_only=c26d1a3e4e0a4506a836972262a9468f].

### Participants

Participants were recruited using non-probability convenience sampling from a larger observational study on distress, inflammatory reactivity, induced SH, and persistent pain in adults with HIV ^[36]^. Although sex and age matching were applied in the larger study, this was relevant only to analyses of inflammatory outcomes and not to the SH analyses reported here. All individuals who expressed interest were screened for eligibility. Pain status was determined at screening, and consecutive eligible participants from each pain group were invited to participate until target enrolment was reached.

We enrolled people with HIV who were virally suppressed (< 50 copies/ml) and reported either persistent pain or no pain. Participants who reported persistent pain had to report pain on most days for more than three months ^[37]^. Participants were eligible if they were aged 18 to 65 years, fluent in English or isiXhosa, and living with HIV, with recent evidence of viral suppression (viral load < 50 copies/ml within the preceding three months).

Exclusion criteria included pregnancy or suspected pregnancy; electrical or metal implants in the forearm to be tested; sensation problems or tattoos on the forearm to be tested; known neurological or cardiovascular conditions; advice from a medical practitioner to avoid stressful situations; and an acute psychiatric condition (substance use disorder, psychosis, or high suicide risk) identified using screening with the Mini International Neuropsychiatric Interview (MINI), with follow-up using the WHO ASSIST, WHO AUDIT, or the full MINI psychosis module, as appropriate ^[36]^. Suicidal ideation and severity were assessed using adapted MINI items, and participants who met criteria for high suicide risk were excluded and referred for care. The community from which participants were recruited has high rates of unemployment (38%) and informal housing (55%) ^[38]^. Unemployment and informal housing have been linked to distress, which aligns with the focus of this study ^[39–41]^.

Participant pain status was determined using questions adapted from the Brief Pain Inventory (BPI) self-report measure ^[42]^, which has been translated and validated in isiXhosa and used in people with HIV in South Africa ^[43]^. The opening statement to the screening questions was: “*Throughout our lives, most of us have had pain from time to time (such as minor headaches, sprains, and toothaches).* This statement was followed by three screening questions on pain frequency and duration: (i) *“Do you have pain other than these kinds of pain today?”* (ii) “*Other than those day-to-day kinds of pain, do you have pain in part of your body on most days?”* (iii) *“Have you had that pain on most days for more than 3 months?”* Participants who answered ‘yes’ to all three questions were included in the ‘pain’ group, whereas those who answered ‘no’ to all the questions were included in the ‘pain-free’ group. Participants who answered ‘yes’ to questions (i) and (ii), but ‘no’ to question (iii) were excluded because they were deemed to have acute pain. Participants who completed the SH procedure were compensated ZAR 150 (~USD 9.72) for their time in addition to the larger study compensation, whereas those who withdrew during the procedure received *pro rata* compensation. Participants and members of the public were not involved in the design, recruitment, or conduct of this study. People living with persistent pain provided guidance on dissemination.

### Procedure

#### Informed consent and participant orientation

Participants had the option to communicate in English or isiXhosa. An assessor screened participants for eligibility and facilitated the written informed consent process, during which participants were informed that their participation was voluntary, with no impact on their clinical care. Participants were free to withdraw without negative consequences. The assessor then administered the battery of self-report questionnaires (including the Hopkins Symptom Checklist-25 and BPI), drew two blood samples (results not reported here), and scheduled the participant’s SH induction session.

A second assessor (LM), who was blinded to the results of the first assessor, conducted the SH induction and related assessments. The assessor settled the participant on a seat facing her across a table, with the designated pain-free arm resting on the table and a touch monitor facing the participant, to one side. Following a standardised script for consistency, the assessor orientated the participant to the study equipment and procedures. Participants were allowed to ask questions, and then verbally confirmed their previously written consent. The assessor marked the participant’s forearm for the electrode location and the radial lines and calibrated the electrical current for each participant (see Calibration of electrical current). The assessor then demonstrated how to use the electronic touch-based VAS, and participants received an opportunity to practice rating each sensory modality. All practice ratings were recorded but excluded from the analysis.

#### Calibration of electrical current

The individual detection threshold for a single electrical stimulus was used to determine the current for HFS and subsequent assessment of primary hyperalgesia (not reported here). We used an adaptive staircase procedure to calibrate the electrical ^[44]^. First, the current was gradually increased from 0 mA in 0.10 mA increments until the participant reported feeling the stimulus. Second, the current was reduced in 0.5 mA increments until the participant no longer felt the stimulus. Third, the current was increased in 0.2 mA increments until the participant could once again perceive the stimulus. The final current detected in the last step represented the participant’s threshold for detecting a single electrical stimulus and was multiplied by 10 and used for the stimulation intensity for both inducing SH and administering single electrical stimuli during sensory testing, as seen in other studies ^[29]^.

#### Baseline tests

Three rounds of baseline sensory testing were completed in approximately six minutes, using five modalities in a fixed order: 128mN, 256mN, single electrical stimulus, brush and VFF (last three not reported here). For pinprick and VFF trials, one rating was recorded for the average of three stimulations, each lasting ~1 second; for the electrical and brush stimuli, a single stimulus was rated. Similar to the established DFNS procedure ^[45; 46]^, temporal summation was assessed once, after the baseline tests and before the HFS briefing script and SH induction, using a 256mN pinprick probe applied ~3 minutes before the SH induction. This paradigm captures temporal summation of punctate mechanical stimulation, primarily mediated by A-delta afferents ^[45–47]^. All tests were applied approximately 1 cm from the cathode, at slightly different locations to limit interference between stimuli.

#### SH induction

Secondary hyperalgesia was induced by applying HFS to a pain-free area of the forearm, using a circular cathode with 10 blunt steel pins against the anterior forearm and an anode around the upper arm. We chose the forearm that had not undergone venepuncture for blood collection in the primary study because venepuncture can cause local sensitivity, which could interfere with SH assessment. If both arms had undergone venepuncture or the venepuncture-spared arm had a contraindication to HFS (e.g., metal implant), SH induction and assessment were conducted on the forearm used for venepuncture seven days after venepuncture. The HFS consisted of five one-second trains at 100 Hz, each followed by a 9-second inter-train interval. The simulation current was set to 10 times the individual’s detection threshold for a single electrical stimulus, which was calibrated for each participant ^[48]^. Electrical stimulation was delivered using a constant current electrical stimulator (DS7A; Digitimer Limited, Hertfordshire, UK) with a maximum voltage of 400V, a pulse width of 2000μs, and a square pulse waveform. The effect of HFS is typically centred around the cathode. The DS7A was operated in constant current mode, with the voltage control set to its maximum (400 V), as recommended by the manufacturer, to support stable current delivery across individuals with varying skin resistance. In this mode, the stimulator dynamically adjusts the output voltage up to this maximum as needed, depending on each participant’s skin impedance, in order to maintain the target current. Although the stimulator can deliver high voltage, the current output was individualised, and due to the small surface area of the blunt-pin cathode, the stimulation remained superficial. This design minimises the likelihood of muscle contraction, as the depth of current penetration is limited. The study assessments were programmed in Affect5 ^[49]^. The participant received five trains of HFS and rated each one on the VAS. Thus, the induction yielded one VAS rating for each of the five HFS trains from each participant.

#### Follow-up tests

A 30-minute waiting time allowed for the development of SH ^[50; 51]^, which typically becomes apparent within 20-30 minutes after induction using HFS ^[27; 52]^. Therefore, we started testing 30 minutes after the first HFS train to capture the peak effect of SH. During the waiting time, participants were provided with magazines containing content unrelated to the study. The surface area of SH was assessed 30, 45, and 60 minutes after the induction (Figure 1). The 5 sensory tests were re-administered 35, 50, and 65 minutes after the induction (Figure 1).

**Figure 1:**
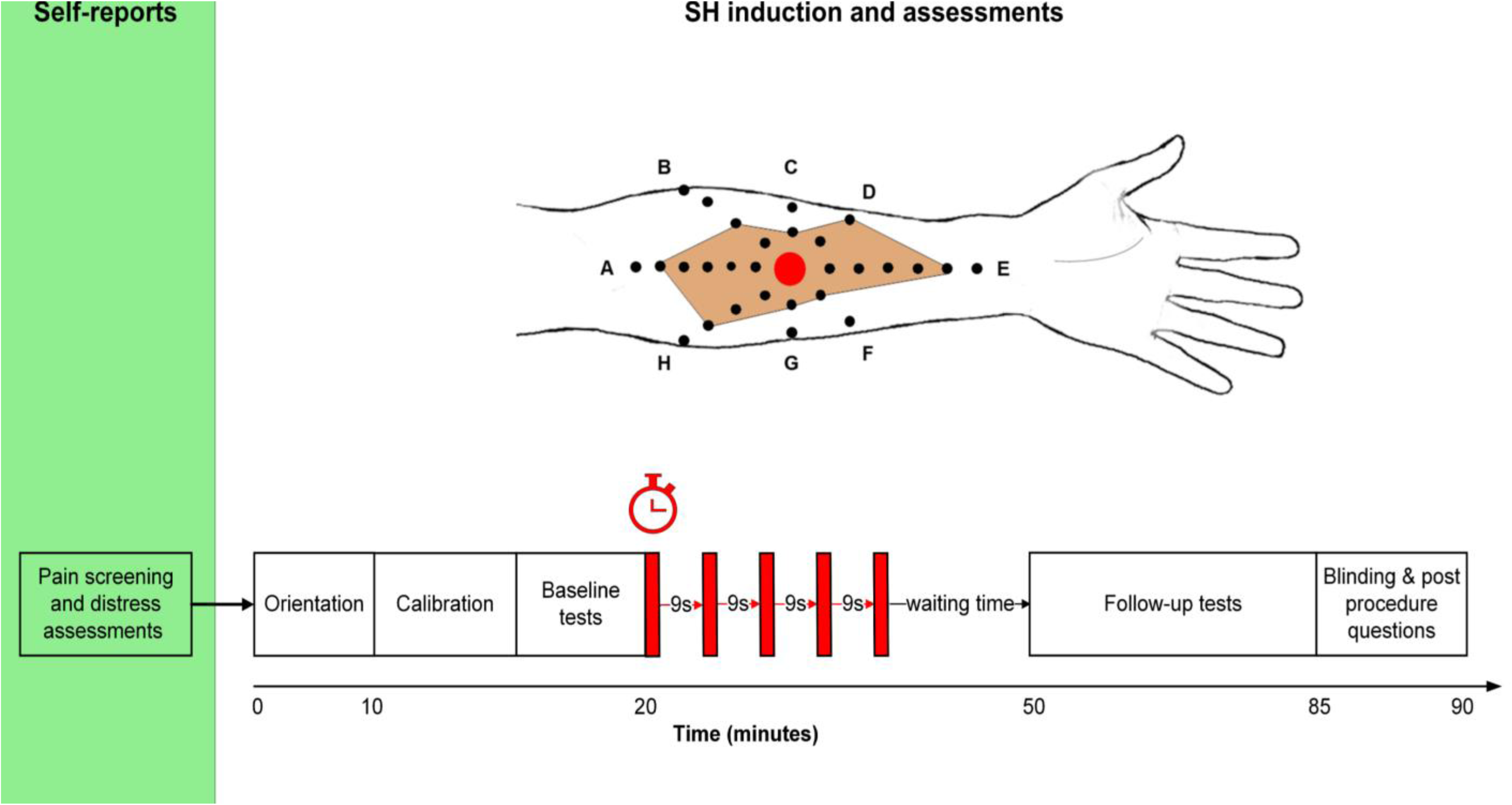
Study procedure. The green box indicates self-reported assessments, and the white box indicates secondary hyperalgesia (SH) induction and assessments. The self-reported assessments were separated from the SH induction and assessments by either a short break or up to 7 days. The five red blocks indicate the high frequency electrical stimulation trains. The red circle centrally located on the radial lines on the forearm indicates the cathode, with each of the 8 lines on the skin spaced 45 degrees apart and the black dots forming the lines 1cm apart (not drawn to scale). The area shaded in orange shows the surface area of SH.

#### Blinding

Two assessors collected study data. The first assessor collected self-reported distress and pain status. The second assessor induced SH using HFS and assessed its surface area and magnitude. Participants were blinded to the study hypotheses; the first assessor was blinded to the study aims and hypotheses; the second assessor was blinded to participant distress self-report and study group (pain or pain-free); the data analyst for hypothesis 2 (people with HIV reporting persistent pain would display greater induced SH than people with HIV reporting no pain) was blinded to the study group.

To support the blinding of participants to the study aims and hypotheses, we withheld information on the study aims and hypotheses, including that participants were being grouped by pain status. Participants completed a blinding assessment after the procedure: they were asked to guess the purpose of the study, and their response was recorded. We applied conservative criteria to assess whether participants remained blinded to the study’s aims and hypotheses.

After the SH assessments, the blinding assessments were completed, first by the assessor, and then by the participant. The assessor had to guess each participant’s study group and rate her confidence in her guess on a five-point Likert scale of “not at all confident”, “not confident”, “confident”, and “extremely confident” (the planned ‘neutral option was omitted by technical error – protocol deviation 1 of 4). Finally, the assessor asked the participant for feedback on her communication and the general experience of the procedure. The second assessor also conducted the preliminary data analysis; therefore, to maintain blinding to the study group, the study data were assigned a second study ID for each participant and the study groups were recoded as ‘a’ and ‘b’ by VJM. The second assessor then conducted the preliminary data analysis and interpretation with this recoding in place, after which the assessor was unblinded to complete the interpretation and write the manuscript.

### Outcomes

#### Independent variable: psychological distress

Psychological distress was assessed using the Hopkins Symptom Checklist-25 (HSCL-25) ^[53]^, which consists of 25 items that estimate symptoms of depression (15 items) and anxiety (10 items). Participants rate how much each symptom (e.g., ‘poor appetite’, ‘feeling lonely’, ‘trembling’, ‘feeling fearful’, etc.) applied to them in the past month on a four-point scale (1 = ‘not at all’ to 4 = ‘extremely’). The final score (and the study measure of distress) was the arithmetic mean of all the item scores and lies between 1.00 and 4.00. The HSCL-25 has been used in South Africa ^[54]^, including with people with HIV ^[55]^. Since the HSCL-25 has not been formally translated and validated in isiXhosa-speaking populations, we used a forward-and-back-translation process to ensure semantic equivalence and cultural relevance to the local population, although we did not perform a formal validation of the translated version ^[56]^.

#### Dependent variable: secondary hyperalgesia

To capture the increase in central nervous system facilitation after a barrage of afferent nociception, we chose a human surrogate model of SH for direct *in vivo* characterisation of the aspects of the response that are thought to rely on central (rather than peripheral) changes ^[29; 57]^. In short, the induction protocol involved the delivery of controlled electrical stimulation to the skin of the forearm (Figure 1). Hyperalgesia to pinprick stimulation of the skin develops in the area surrounding the induction stimulus within 20-30 minutes after the induction. We quantified SH using two measures: surface area (primary measure) and magnitude (secondary measure).

##### Primary measure of SH: surface area

The surface area of SH was estimated using the 8-radial-lines method ^[58; 59]^, which identifies the boundary between hypersensitive and normosensitive skin by using repeated stimuli along 8 radial lines that transect the site of stimulation at 45° angles. We used the method previously reported in ^[60]^, except that we used a 128mN Von Frey filament (Marstock nervtest, Germany), as in ^[61]^. The surface area was assessed 30, 45, and 60 minutes after the induction.

##### Secondary outcome: magnitude of SH

Participants gave Visual Analogue Scale (VAS) ratings of pain to two punctate mechanical stimuli (128 mN and 256 mN; PINPRICK, MRC systems, Heidelberg, Germany) applied for ~1 s, at three different points within 1 cm of the electrode. The scale was a touch-based, electronic, vertical VAS, anchored with 0 for “no pain” at the bottom and 100 for “pain as bad as you can imagine” at the top (anchors translated for isiXhosa-speaking participants). Anchors were translated using a forward-and-back-translation process to ensure semantic equivalence and local relevance and no formal psychometric validation was undertaken. Participants selected their rating by swiping a stylus pen across the vertical scale on the screen, prompting the bar to fill up with red to the level of the rating. This rating was recorded electronically as a number between 0 and 100. The vertical VAS is a valid and reliable measure that is easy to use, requires less reliance on numeracy concepts and is suitable for different adult populations ^[62–65]^. This assessment was performed three times immediately before the induction and at 35, 50, and 65 minutes after the induction.

#### Exploratory variable

Recognising that high frequency electrical stimulation (HFS) is commonly reported to be painful and that the relationship between HFS painfulness and SH is controversial ^[52]^, we assessed the painfulness of the induction trains. Participants rated each train of HFS using the electronic VAS. The results of this assessment were reported descriptively, and we planned to include this variable as a covariate.

### Data handling and analysis

Participant demographic information, distress and pain status data were recorded using the University of Cape Town’s RedCap database ^[66; 67]^. The data on SH outcomes were directly recorded into Affect5, and additional procedural notes were manually recorded and later transcribed into an Excel sheet. All data from RedCap, Affect5, and Excel were imported into R for analysis, using R version 4.2.0 (The R Foundation for Statistical Computing) and RStudio (Integrated Development Environment for R 2023 version 12.1.402) ^[68]^. The packages used were: tidyverse ^[69]^, readxl ^[70]^, gridExtra ^[71]^, here ^[72]^, kableExtra ^[73]^, ggstatsplot ^[74]^, pracma ^[75]^, dplyr ^[76]^, readr ^[77]^, arsenal ^[78]^, bayestestR ^[79]^, DescTools ^[80]^,rcompanion ^[81]^, performance ^[82]^, ggrepel ^[83]^, formatR ^[84]^, magrittr ^[85]^, ggeffects ^[86]^, lme4 ^[87]^,and rlmer ^[88]^.

The target sample size was determined pragmatically: given the sourcing of participants from the pool of ~100 participants in the larger study, the more restrictive inclusion criteria for the SH induction, and the likelihood of some attrition. We planned to recruit 60 participants (30/group) to the SH induction. This decision was guided by both feasibility constraints (time, participant availability) and statistical considerations. Previous studies of experimentally induced SH using HFS without between-group comparisons had used samples of 7-20 ^[29; 51]^, and we deemed the 85% power to detect a correlation of r=0.40 (when alpha is 0.05) offered by a sample size of n=53 to be sufficient. We acknowledge that the cited effect sizes were derived from homogeneous samples of healthy young adults, whereas our sample included more variability due to HIV, persistent pain, psychological distress, and socioeconomic adversity. As such, these estimates may not fully reflect the variability within our study population. To contextualise the statistical power provided by our final sample, a sample size of n=45 would be expected to provide 80% power to detect a correlation of r=0.41 when alpha is 0.05.

Demographic data were presented in tables and reported descriptively. Frequencies and proportions are reported for categorical variables and median (IQR) for numerical variables. In all box-and-whisker plots, individual participant scores are represented by dots, while horizontal lines show the 25th, 50th (median), and 75th percentiles. The whiskers of the boxplots indicate the spread of the data beyond the IQR: the upper whisker extends from Q3 to reach the maximum data point that falls within 1.5 times the IQR, whereas the lower whisker extends from Q1 to reach the minimum data point that falls within 1.5 times the IQR.

#### Primary and secondary measures

Both hypotheses were tested together, using one model for each measure of SH. The dependent variable was the surface area (primary measure) or magnitude (secondary measure) of SH.

For surface area, the within-participant area was the sum of the areas of the 8 triangles formed by the transition points on the radial lines, resulting in one estimate of surface area for each of the three post-induction time points. In the protocol, we planned to sum the area across the three time points for each participant, using the ‘area under the line’ for both surface area and magnitude of SH. However, given that the area under the line can yield unreliable estimates when few replicates are available, we opted to include all three assessments, clustered as repeated measurements within each participant, in a linear mixed model in R (protocol deviation 2 of 4).

For magnitude, the mean ratings for the two punctate mechanical weights were calculated for each time point, and the mean rating before induction was subtracted from the mean rating at each post-induction time point. This was protocol deviation 3 of 4: in the protocol, we had planned to express follow-up ratings as a percentage of baseline mean ratings, but we opted for the difference calculation to avoid artificially inflated statistical estimates of effect.

The independent variables were distress and group. A random factor allowed a different intercept for each individual; the three repeated measures were nested within each individual. Unadjusted models were specified, followed by models adjusted by three covariates: the current used for SH induction (calibrated to individual), the within-participant median of the ratings of the HFS induction trains, and the number of days between distress self-report and SH induction. The last covariate was relevant to only a few participants (n=8) but was included in case the delay led to the distress score poorly representing the participant’s state at the time of SH induction and assessment. For only the magnitude models, the within-participant mean of all pinprick ratings (128 mN and 256 mN) for the baseline timepoint was also included as a covariate, to control for baseline differences in sensitivity to pinprick stimulation.

We visualised model assumptions by generating plots to check the following model assumptions: normality of residuals, linearity, homogeneity of variance, influential observations, and collinearity, where applicable. Given that the conventional linear mixed models violated model assumptions, we conducted robust linear regression to account for influential observations. Next, we computed bootstrapped 95% confidence intervals for the robust regression estimates to account for the violations of the normality assumption.

Bootstrapping provides a way to assess the stability of the findings by resampling the data and evaluating the variability of the regression estimates across multiple samples to offer a more reliable estimate of model uncertainty ^[89]^. We report effects as estimates bootstrapped from covariate-adjusted robust models including model parameters without bootstrapping. We opted against transforming the data given the nature of the violations of distributional assumptions and to retain interpretability

#### Blinding analysis

We report the percentage of participants who correctly guessed the aim of the study. We also report the percentage of participants for whom the second assessor correctly guessed group membership. Given that no single method perfectly captures blinding effectiveness, we assessed the blinding of the second assessor using three methods.

First, we used Cohen’s Kappa statistic (protocol deviation 4 of 4). Whereas James’s Blinding Index, which was planned in the protocol, prioritises a “do not know” response that was not offered in our design ^[90]^, Cohen’s Kappa is suitable for a two-category forced-choice design like ours (“pain” or “pain-free”) ^[91]^. Second, we used Chi-square goodness-of-fit tests to assess whether the observed distribution of guesses about group membership differed from the expected distribution based on a 50% chance of guessing correctly (which would reflect random guessing and retained blinding) ^[92]^. Third, we also considered the match between correct guesses about group membership and assessor-reported confidence. The availability of confidence ratings allowed us to look more closely at potential unblinding for a subgroup of participants about whose group status the assessor reported feeling confident. Therefore, we drew the data on only those participants for whom the assessor reported confidence of 4 or 5 and conducted a separate chi-square test on the group guess data, comparing the actual frequency of accurate group guesses to the 50% frequency expected under chance conditions (i.e. random guessing; blinding retained). Finally, we repeated the main analyses without participants for whom the second assessor had accurately and confidently guessed group membership, as identified above, to assess the sensitivity of the main findings to potentially broken blinding.

#### Planned exploratory analyses

We plotted the VAS ratings captured for the five HFS trains and reported these data descriptively. Due to a technical problem, approximately 49% of VAS ratings for HFS trains were missing. To ascertain the reason for missingness in the context of our sample size (n=45), we investigated the missing data patterns visually and descriptively and found no systematic differences in missingness (Figure S1). However, given that ratings of HFS are known to increase across the 5 trains and to allow for group differences, missing values were imputed using the within-group median rating of each HFS train (1–5), as calculated from the available data. The main regression analyses included HFS train ratings as a covariate; imputed data were used for these. Given the quantity of imputed HFS train ratings, we also constructed a second set of adjusted models without HFS train ratings as a covariate.

#### Post-hoc exploratory analysis

We visually investigated whether withdrawal from the procedure was predicted by distress severity, and planned to follow up any visual indication of a relationship using logistic regression. We planned to follow up on any significant result with interaction plots ^[93]^.

## Results

Data were collected from 9 February 2021 to 24 November 2021.

### Participants

Fifty-six participants, 16 identifying as males and 40 as females, were deemed eligible and enrolled in the study. Five participants withdrew during the procedure; three did not complete the procedure due to technical error; three were identified as ineligible upon demographic data inspection (details in Figure 2). Complete datasets were available for 45 eligible participants (Table 1, pain: n=19; pain-free: n=26). None of the participants reported taking analgesic medication within 24 hours before the induction. For pain *in the past week*, low back pain was the most reported site (n=11) and the most common site of worst pain (n=10). For pain *for the past 3 months*, upper back pain was the most reported site (n=11), whereas the upper back (n=9) was reported as frequently as the low back (n=9) as the site of the worst pain. Data on pain severity (for *pain in the past week* and *pain for the past 3 months*) and interference for all *pain in the past week*, presented only to participants reporting persistent pain are shown in Table 1. The baseline ratings to pinprick stimulation were higher in the pain group (7.50(0.67-22.67)) than in the pain-free group (1.00(0.33-3.38), p=0.05).

**Figure 2:**
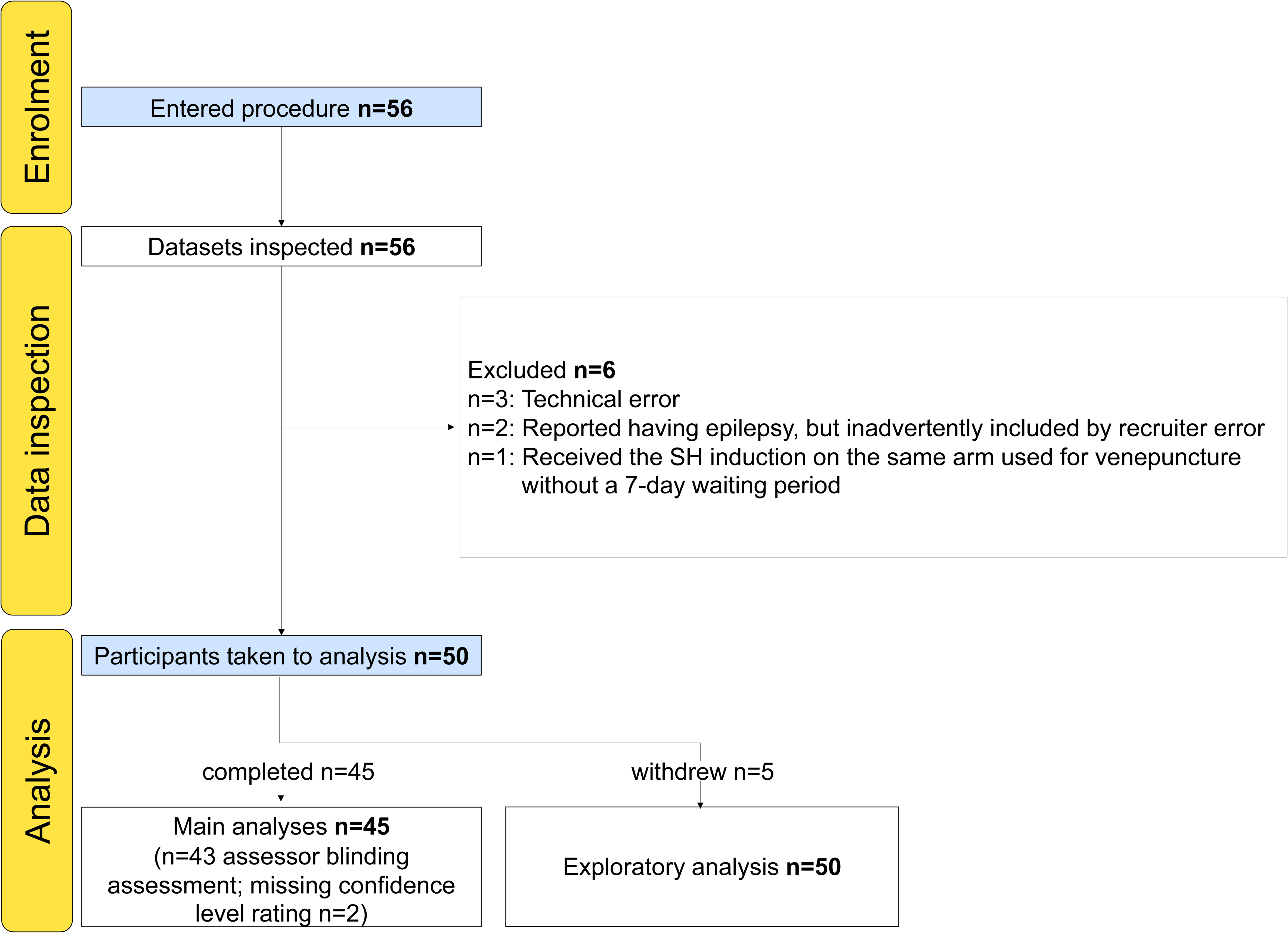
Flowchart of participants enrolled and taken to the analysis.

**Table 1:** Descriptive data (n=45). The group with persistent pain reported higher distress severity, and higher baseline ratings to pinprick stimulation (not reported) than the pain-free group. After imputation, between-group comparison of ratings of HFS trains showed a difference that was not present in the data before imputation. No other significant differences were noted between the groups. HSCL-25: Hopkins 25-item symptom checklist

|  | Pain (N=19) | Pain-free (N=26) | Total (N=45) | p-value |
| --- | --- | --- | --- | --- |
| **Age (years)** |  |  |  | 0.765 |
| Median | 43.00 | 41.00 | 41.00 |  |
| Q1, Q3 | 36.00, 48.50 | 34.25, 46.75 | 35.00, 47.00 |  |
| Range | 30.00 - 58.00 | 31.00 - 64.00 | 30.00 - 64.00 |  |
| Mean (95% CI) | 43.11 | 42.92 | 43.00 |  |
| SD | 8.65 | 9.62 | 9.12 |  |
| **Sex (n (%))** |  |  |  | 0.478 |
| female | 12 (63.2%) | 19 (73.1%) | 31 (68.9%) |  |
| male | 7 (36.8%) | 7 (26.9%) | 14 (31.1%) |  |
| **Current used for HFS (mA)** |  |  |  | 0.853 |
| Median | 0.12 | 0.16 | 0.14 |  |
| Q1, Q3 | 0.08, 0.19 | 0.09, 0.17 | 0.09, 0.18 |  |
| Range | 0.04 - 0.27 | 0.04 - 0.26 | 0.04 - 0.27 |  |
| Mean (95% CI) | 0.14 | 0.14 | 0.14 |  |
| SD | 0.07 | 0.06 | 0.06 |  |
| **Distress severity HSCL-25: mean score 1-4)** |  |  |  | < 0.001 |
| Median | 2.24 | 1.38 | 1.48 |  |
| Q1, Q3 | 1.68, 2.58 | 1.08, 1.60 | 1.12, 2.24 |  |
| Range | 1.08 - 3.36 | 1.00 - 2.52 | 1.00 - 3.36 |  |
| Mean (95% CI) | 2.14 | 1.46 | 1.75 |  |
| SD | 0.64 | 0.47 | 0.64 |  |
| **Pain severity for pain in the past week (BPI mean score 0-10)** |  |  |  |  |
| Median | 5.25 | NA | 5.25 |  |
| Q1, Q3 | 4.12, 6.00 | NA | 4.12, 6.00 |  |
| Range | 3.00 - 7.50 | NA | 3.00 - 7.50 |  |
| Mean (95% CI) | 5.13 | NA | 5.13 |  |
| SD | 1.30 | NA | 1.30 |  |
| **Pain severity for pain for the past 3 months (BPI mean score 0-10)** |  |  |  |  |
| Median | 5.00 | NA | 5.00 |  |

|  | Pain (N=19) | Pain-free<br>(N=26) | Total (N=45) | p-value |
| --- | --- | --- | --- | --- |
| Q1, Q3 | 4.00, 5.88 | NA | 4.00, 5.88 |  |
| Range | 3.25 - 8.25 | NA | 3.25 - 8.25 |  |
| Mean (95% CI) | 5.21 | NA | 5.21 |  |
| SD | 1.53 | NA | 1.53 |  |
| **Pain interference for all**<br>**pain in the past week (BPI**<br>**mean score 0-10)** |  |  |  |  |
| Median | 5.00 | NA | 5.00 |  |
| Q1, Q3 | 3.93, 6.14 | NA | 3.93, 6.14 |  |
| Range | 3.00 - 9.43 | NA | 3.00 - 9.43 |  |
| Mean (95% CI) | 5.39 | NA | 5.39 |  |
| SD | 1.93 | NA | 1.93 |  |

### Results of HFS induction

The median (IQR) current used for HFS was 0.14 mA (0.09-0.18 mA) and there was no difference in current between groups (Table 1, p=0.9). Nearly half (126 of 255, 49%; pain: 50; pain-free: 73) of the VAS ratings of the HFS trains were missing due to a technical problem. We used the available data to calculate the median (IQR) ratings of the HFS trains for the sample and found 48 (10-82) on a VAS expressed from 0-100. There was no difference in the ratings of HFS trains between the pain (61(13-90)) and pain-free (44(8-73)) groups (p=0.13), but the group-level ratings differed between trains and were non-parametrically distributed (Figure S2). Therefore, for the adjusted models in the main analyses, we imputed the missing ratings for HFS trains by using the group-specific median rating for each train. When the imputed ratings were excluded from the second set of adjusted models, the pattern, direction, and magnitude of effects were unchanged (Table S1).

The median (IQR) surface area of SH for the sample was 22.27cm^2^ (5.66-49.85). Of the 135 surface area data points (three per participant), 12 participants had no SH at T30, eight had no SH at T45, and 10 had no SH at T60. Five (of 45) participants showed hyposensitivity at all post-induction time points. The median (IQR) magnitude of SH for the sample was 1.17 (0-7.67). Of the 135 post-induction data points, hyposensitivity was observed at 21 data points at T35, 22 at T50, and 23 at T65. Eight (of 45) participants showed hyposensitivity at all time points. Both the surface area (p=0.15) and magnitude (p=0.61) of SH were no different between groups.

### Participants with persistent pain reported more distress

The median (IQR) distress severity for the sample was 1.48 (1.12-2.24). The pain group reported significantly higher distress severity (2.24 (1.68-2.58)) than the pain-free group (1.38 (1.08-1.60)) (p<0.001, Table 1).

### Distress positively predicted induced SH area and magnitude

Figure 3A-D shows the relationship between distress and the surface area (A, B) and magnitude (C, D) of SH without accounting for the clustered nature of the data. Formal analysis showed that distress severity was positively associated with the surface area of SH in both unadjusted and covariate-adjusted models across conventional (Table S2, model diagnostics Figures S3 and S4) and robust models (Table S3). On average, a 1-unit increase in distress was associated with an average 19.10 cm^2^ (Table 2, 95% CI: 2.85-35.34; p=0.02) increase in the surface area of SH, with all other variables held constant.

**Figure 3:**
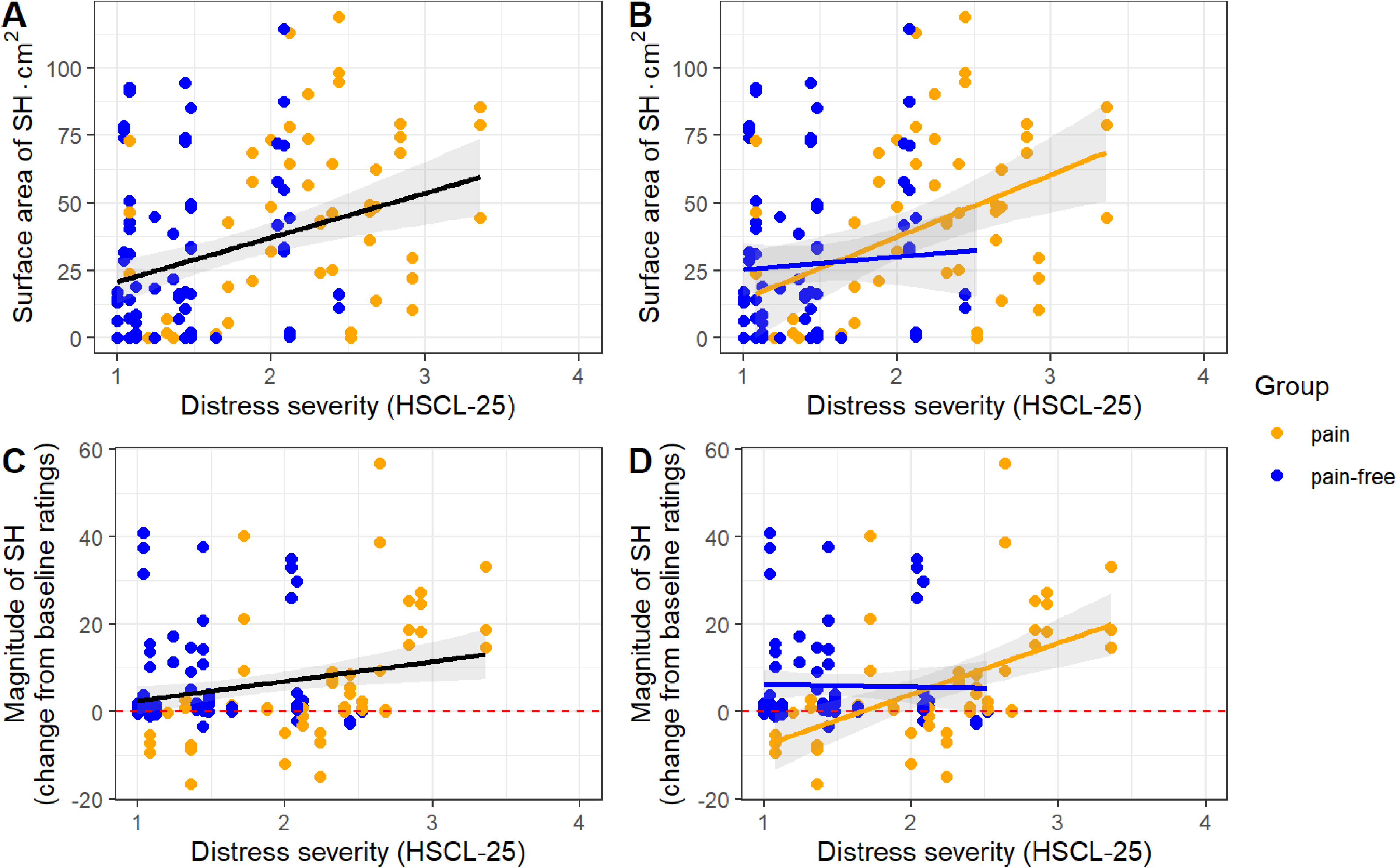
Scatterplots (A-D) overlain with the best-fitting straight line (ribbon = 95% CI), not the regression model. Each dot represents a participant’s score at one of three time points. Panels A and B show relationships between distress and secondary hyperalgesia (SH) surface area for the whole sample (n = 45; primary hypothesis) and stratified by group, respectively. Panels C and D show relationships between distress and SH magnitude for the whole sample and stratified by group, respectively. Magnitude is expressed as the change in rating from the mean of the three baseline assessments to each follow-up assessment. The horizontal red dotted line indicates no SH.

**Table 2:** Bootstrapped covariate-adjusted robust models predicting surface area and magnitude of secondary hyperalgesia (SH). pid: individual participant code.

| *Predictors* | **Surface area of SH** |  | **Magnitude of SH** |  |
| --- | --- | --- | --- | --- |
|  | *Estimates* | *CI* | *Estimates* | *CI* |
| Intercept | 3.98 | -30.91 – 38.87 | -3.42 | -13.93 – 7.09 |
| Distress severity | 19.10 * | 2.85 – 35.34 | 6.24 * | 1.28 – 11.21 |
| Group (pain) | -1.88 | -24.03 – 20.27 | -6.92 * | -13.70 – -0.13 |
| Current used for HFS | -117.83 | -257.02 – 21.36 | -29.36 | -71.42 – 12.71 |
| HFS painfulness | 0.13 | -0.25 – 0.51 | 0.03 | -0.10 – 0.17 |
| Time difference<br>between distress &<br>SH assessments | 1.07 | -1.06 – 3.20 | 0.05 | -0.60 – 0.71 |
| Baseline mean<br>pinprick ratings |  |  | 0.25 * | 0.02 – 0.48 |
| **Random Effects** |  |  |  |  |
| $\sigma^2$ | 178.63 | | 2.99 | |
| $\tau_{00}$ | 737.41<sub>pid</sub> | | 71.19<sub>pid</sub> | |
| ICC | 0.81 |  | 0.96 |  |
| N | 45<sub>pid</sub> |  | 45<sub>pid</sub> |  |
| Observations | 135 |  | 135 |  |
| Marginal R<sup>2</sup> /<br>Conditional R<sup>2</sup> | 0.192 / 0.842 |  | 0.240 / 0.969 |  |

\* $p < 0.05$ \*\* $p < 0.01$ \*\*\* $p < 0.001$

For the magnitude of SH, distress severity was positively associated with the magnitude of SH in all models except the unadjusted conventional model (Table S2-S3, model diagnostics Figures S5 and S6). On average, a 1-unit increase in distress was associated with an average increase in the magnitude of SH of 6.24 (Table 2, 95% CI: 1.28-11.21; p=0.01) (change in rating on a 0-100 scale).

### Only the magnitude of induced SH was related to pain status

Figure 4A-B shows the surface area (panel A) and magnitude (panel B) of SH over time between groups. For the surface area of SH, the main effect of group was not statistically significant (p=0.87). For the magnitude of SH, the main effect of group was statistically significant: on average, magnitude was marginally *lower* in participants with pain than in pain-free participants, by 6.92 units (95% CI: −13.70 to −0.13, p = 0.05).

**Figure 4:**
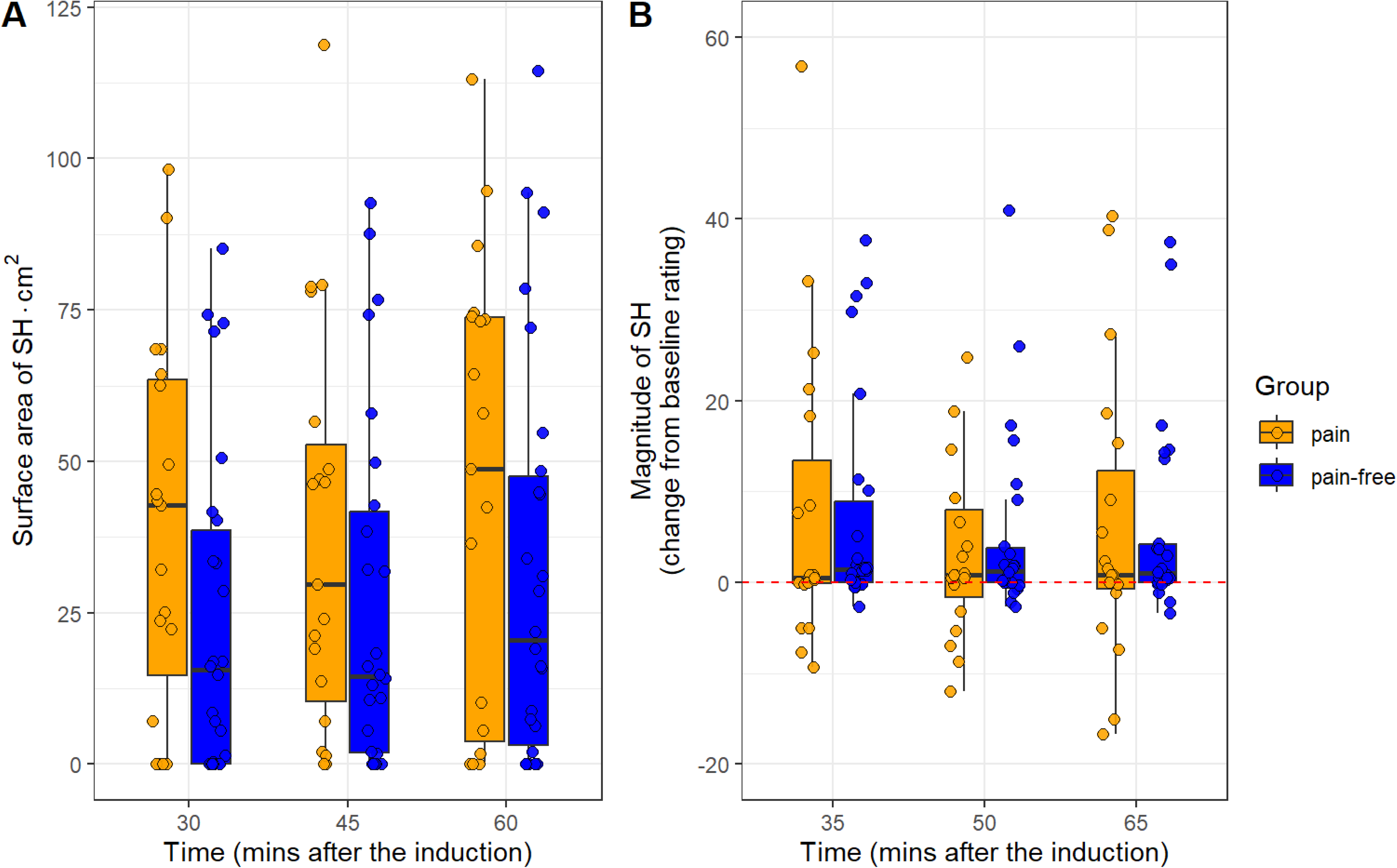
The surface area (A) and magnitude (B) of secondary hyperalgesia (SH) over time. Each dot represents a participant’s score at one of three time points. Magnitude is expressed as the change in rating from the mean of the three baseline assessments to each follow-up assessment. The horizontal red dotted line indicates no SH.

### People with pain showed a stronger relationship between distress and SH magnitude

Given that Figures 3B and 3D suggested that the strength of the associations between distress and the two SH measures might differ between groups, we included an exploratory analysis of the interaction between distress and group in each covariate-adjusted robust regression model (Table S4). For the surface area of SH, there was no interaction between distress and group (Table S4, bootstrapped coefficient: 17.63; 95% CI: −14.48-49.75; p=0.28). For the magnitude of SH, there was a significant interaction between magnitude and group (Table S4, bootstrapped coefficient: 13.13; 95% CI: 4.04-22.21; p<0.01): the relationship between distress and magnitude was greater in the group with pain than in the pain-free group (Figure 3D).

### Blinding check

None of the participants correctly guessed the aim of the study. The assessor correctly guessed the group membership of 20 out of 45 participants (44%). Cohen’s Kappa estimate was −0.13 (95% CI: −0.42-0.16), indicating no meaningful agreement between the assessor’s guesses and the actual group memberships. The Chi-square goodness-of-fit tests confirmed no relationship between the actual group and the assessor’s guess of the group (pain: X²=1.3; p=0.3; pain-free: X²=0.03, p=0.8). There were 4 instances of correct group guesses with confidence rated 4 or 5 (pain: n=2; pain-free: n=2), and 11 instances of incorrect group guesses with confidence rated 4 or 5 (pain: n=6; pain-free: n=5). The Chi-square goodness-of-fit tests on these data confirmed no relationship between the actual group and the assessor’s guess of the group (pain: X²=2; p=0.2; pain-free: X²=0.5, p=0.5). Together, these assessments suggest that participant and assessor blinding was maintained throughout the procedure. Nevertheless, as planned, we excluded instances where blinding might have been compromised (n=4) and reran the main analyses. The main effects of distress on the surface area (p=0.03) and magnitude (p=0.04) of SH remained significant. However, the between-group difference in magnitude of SH was no longer statistically significant (p=0.30).

### Post-hoc exploratory analysis

There was no significant difference in distress between participants who withdrew (n=5, median HSCL-25 score: 1.44; IQR: 1.20-1.80) and those who completed the study procedure (n=45, median: 1.48; IQR: 1.12-2.24; p=0.6).

## Discussion

The current study aimed to clarify the relationship of induced secondary hyperalgesia (SH) to symptoms of depression and anxiety—here, ‘psychological distress’—and persistent pain status, in people with HIV. The primary hypothesis was that symptoms of depression and anxiety would be positively associated with induced SH. This hypothesis was upheld: distress was positively associated with both the surface area and the magnitude of induced SH. The secondary hypothesis was that individuals reporting persistent pain would display greater induced SH than those reporting no pain. This hypothesis was not upheld: participants with persistent pain showed no difference in the surface area of SH compared to pain-free participants, and although those with pain displayed a marginally lower magnitude of SH, we interpret this relationship as possibly spurious, because there was no evidence to reject the null hypothesis once participants with potentially compromised blinding (based on accurate and confident assessor guesses) had been excluded from the analysis.

The strength of the relationship between symptoms of depression and anxiety and induced SH is noteworthy. To support interpretation of the effect size, figure 5A-B visualises the average increase in SH surface area that was associated with a 1-unit increase in distress, at the extremes observed in this study (0-19cm² in panel A and 99-118cm² in panel B). A 6-unit increase in SH magnitude per 1-unit change in distress may not be meaningful when considering the common smallest threshold for a clinically significant change in pain rating of ~20mm on a 100mm scale ^[94]^. However, HFS likely falls short of modelling the total nociceptive load initiated by clinical tissue injury, given that HFS does not recruit the peripheral sensitisation that would be recruited by actual tissue injury ^[29; 95]^. Unlike clinical SH, experimental SH is short-lived and the experimental context of HFS also lacks other cues for threat value that would likely enhance signalling in real-life scenarios of tissue injury ^[60]^. Therefore, under real-life conditions, symptoms of depression and anxiety may support a greater increase in central neuronal responsiveness than that observed in this study, although this possibility remains to be tested.

**Figure 5:**
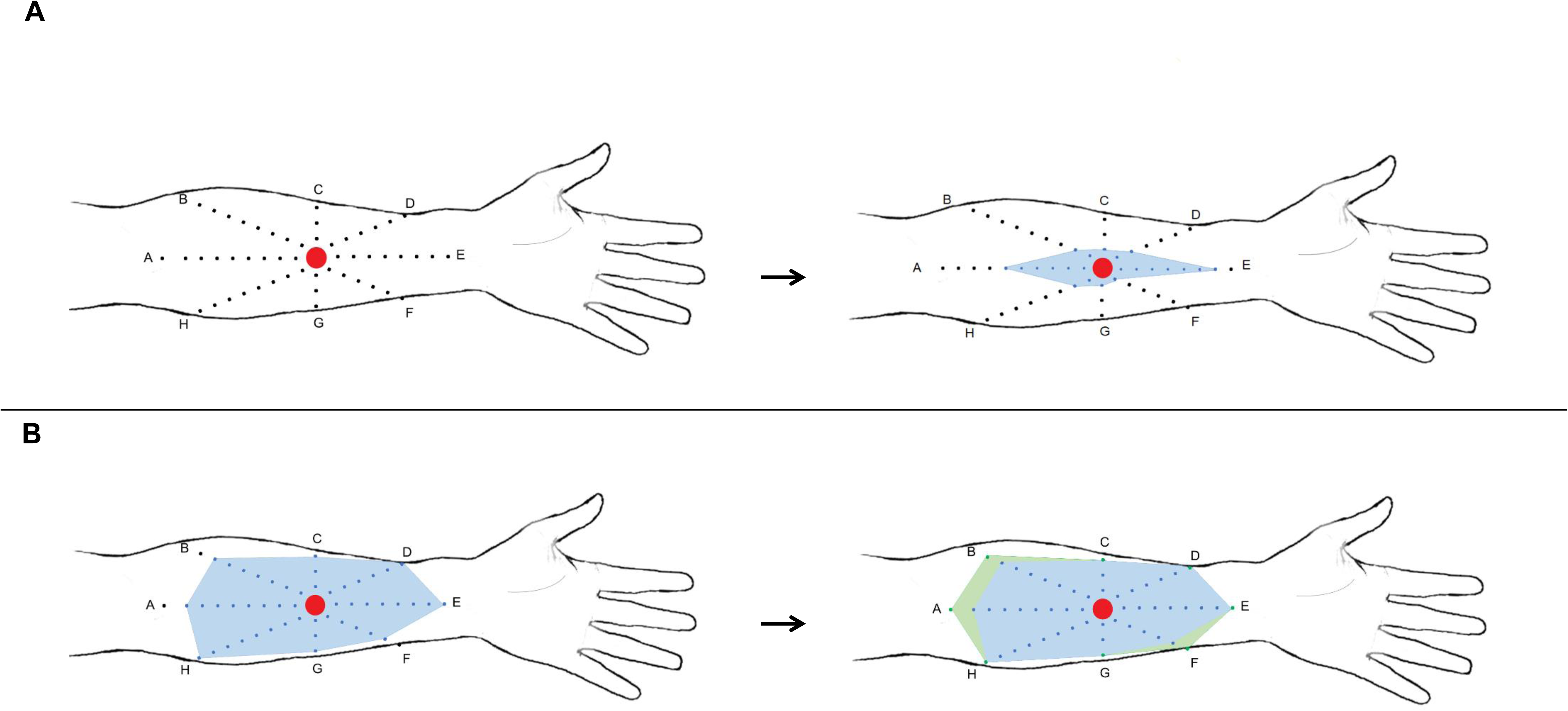
Represents 19cm^2^ increases at the two extremes of surface area observed, to illustrate the estimated effect of a 1-point increase in distress on surface area secondary hyperalgesia (SH). (A) shows a difference between 0 and 19cm^2^ (left to right); (B) shows a difference between 99 (blue) and 118cm^2^ (green). The areas shaded in blue and green show the mapped surface area of SH. Coloured dots mark areas with distinct changes in sensation, and the red circle indicates the cathode’s position. Figure not drawn to scale.

Our results are in alignment with previous work showing that psychological distress and distress-related factors, such as negative affect, can influence neuronal hyperresponsiveness to pain. Interventions to reduce negative affect also reduce SH: in women with a history of trauma, emotional disclosure reduced both negative affect and SH one month later ^[96]^. Similarly, cognitive training to reduce pain catastrophising, and social support from a romantic partner, have each been shown to reduce induced SH on certain measures ^[97–99]^. Our findings extend this literature by showing that current symptoms of depression and anxiety (that, in this study, may be independent of pain) are associated with greater induced SH. Although this hints at a potential influence of psychological distress, as a broader affective state, on central neuronal responsiveness to somatosensory stimuli, it does not confirm causality, because distress could arise secondary to or in parallel with nociceptive responsiveness. Nonetheless, our findings hint that it could be prudent to address symptoms of depression and anxiety alongside pain when they occur clinically, for maximum benefit.

Our secondary hypothesis was that participants with persistent pain would show greater induced central neuronal hyperresponsiveness than participants without pain. Surprisingly, our data did not support this, despite reports of central neuronal hyperresponsiveness in other persistent pain conditions, including fibromyalgia ^[100; 101]^, chronic whiplash ^[102]^, unilateral shoulder pain ^[103]^, chronic pelvic pain ^[104]^, rheumatoid arthritis ^[105]^, and osteoarthritis ^[106]^. Spinal long-term potentiation is one of the mechanisms proposed for these findings (alongside impaired descending inhibitory control) ^[29; 48; 107]^. HFS has been shown to induce similar behavioural changes in humans and rodents, and to cause spinal long-term potentiation-like changes in the rodents ^[28]^. Although this evidence supports the suitability of the HFS model, we did not find more HFS-induced SH in people with persistent pain than in people with no pain. Previous work on central neuronal hyperresponsiveness in people with HIV is limited to homotopic sensitisation and has demonstrated links between temporal summation of painful stimuli and either viral suppression ^[21]^ or persistent pain status ^[22]^ – although the latter was found in a predominantly male sample with mixed levels of viral suppression. Homotopic and heterotopic processes seem to rely on different substrates, as shown by the lack of association between temporal summation and the area or magnitude of HFS-induced SH in healthy humans ^[28]^. The current findings suggest that heterotopic spinal LTP-like processes are unlikely to distinguish between people with HIV based on persistent pain status, and suggest that future comparisons of central neuronal hyperresponsiveness between people with and without persistent pain should control for distress as a potential confounder.

There are three possible alternative explanations for our finding that participants with HIV and persistent pain did not display greater neuronal responsiveness than pain-free participants. The first possibility is that differences in neuronal responsiveness between people with and without persistent pain may reflect differences in inhibition. However, there is evidence that the response to high-frequency electrical stimulation elicits both facilitatory and inhibitory activity, although inhibitory processes were not the primary focus of the current study ^[28]^. The second possibility is that neuronal hyperresponsiveness is somatotopically localised to synapses receiving information from the specific body site(s) where clinical pain occurs ^[108]^. Because we induced SH at a standardised, pain-free site to detect globally increased neuronal responsiveness such as that commonly hypothesised to underpin widespread pain ^[109]^, we would have missed any somatotopically localised hyperresponsiveness. However, a recent study relating susceptibility to induced SH to post-thoracotomy pain also used the forearm as an induction site and did confirm a relationship, suggesting hyperresponsiveness was not somatotopically localised – although the analysis did not control for distress ^[31]^. The third possibility is that our between-group comparison of SH may have been underpowered to detect a difference because our sample size was pragmatically determined and our power calculations were undertaken for the primary analysis.

Our finding that symptoms of depression and anxiety are positively associated with induced SH requires replication in other groups and could be specific to people with virally suppressed HIV, although this seems unlikely in light of the known links between distress and pain in other groups ^[110–112]^. Future research could advance this line of inquiry by investigating whether symptoms of depression and anxiety increase vulnerability to excessive pain after real-life noxious events. For example, it seems plausible that the distress-targeting interventions (e.g., cognitive-behavioural therapy for pre-surgical use) and routine first-line therapies for post-injury use (e.g. pain science education) may also reduce neuronal hyperresponsiveness ^[113]^– although importantly, this remains to be tested, and the general benefits of pre-surgical cognitive behavioural therapy are uncertain ^[114; 115]^. Indeed, the limited benefits of directly targeting SH reinforce the need to identify and target upstream contributors to SH, ideally before or immediately after the noxious insult itself ^[116]^.

The current study has important limitations. First, our approach of screening for the surface area of SH – motivated by the typically greater spread along the proximal-distal axis than the medial-lateral axis ^[117]^ and risk of overestimating without screening ^[60]^– could have missed atypically distributed SH. Second, nearly half of the VAS ratings for the HFS trains were missing and imputed using within-group medians so that this covariate could be included in the main regression models. Although removing the HFS covariate did not alter the overall nature or direction of relationships, the high proportion of missing data may have reduced the precision of the effect size estimates. Accordingly, the effect size estimates themselves should be interpreted with some caution ^[118]^. Third, the study was conducted in people with HIV, and therefore needs replication in the general population. Although HIV is so prevalent in South Africa that people with HIV are not easily distinguished and are reasonably representative of the general population, they would differ in HIV-specific physiology ^[26; 119; 120]^. Fourth, our ethically motivated exclusion of individuals with acute psychiatric conditions, including substance use disorder, psychosis, and suicide risk may have limited our ability to estimate associations at higher levels of psychological distress. Fifth, we assessed neither state distress nor participants’ pain on the day of testing; we only confirmed the absence of pain in the forearm to be tested. Future studies could capture this information to appraise whether state distress is altered by the experimental procedure and whether state distress or day-of-assessment pain influences induced SH. The isiXhosa HSCL-25 and VAS were not formally validated. Finally, our modest sample size may have limited our ability to detect effects in this heterogeneous clinical population, particularly in the secondary analysis.

An important strength of this study is that we induced SH in a clinical population, not just pain-free controls, which is an important translational step towards clarifying the clinical relevance of induced SH. The execution of this study also enacted principles of transparent science: we published the study protocol, locked the analysis plan, reported deviations from the protocol, made analysis scripts publicly available, and openly discussed model assumptions and actions taken to address any violations.

## Conclusion

In this study, symptoms of depression and anxiety were positively associated with both the surface area and magnitude of induced SH in people with suppressed HIV, and this was consistent across participants with persistent pain and participants with no pain. Alongside the evidence that heightened SH reflects enhanced nociceptive responsiveness and arguments that heightened nociceptive responsiveness may predispose people to problematic pain, this work provides a mechanistic measure that could be useful in trials of interventions that address symptoms of depression and anxiety in people approaching painful events, to improve pain outcomes. Although there is little basis to think this finding is specific to people with HIV, its relevance to people without HIV is yet to be tested.

## Data Availability

The de-identified study data are available for selective sharing, subject to review of individual data requests, the use of secure computer platforms, formal use agreements, and compliance with the institutional human research ethics policies. Data use is limited to research purposes, on secure computer servers. The principal investigator (VJM) is the contact person for requests to share data.

https://osf.io/2hdpy/?view_only=c26d1a3e4e0a4506a836972262a9468f

## Acknowledgements

We thank Mathijs Franssen for research support, Kessie Govender for making the electrodes, Nomvula Mdwaba and Andiswa Siyoko for recruiting participants, and Andiswa Gidana and Yoliswa Mtingeni for collecting self-reported data. We are grateful to those who participated in this study. The de-identified study data are available for selective sharing, subject to review of individual data requests, the use of secure computer platforms, formal use agreements, and compliance with the institutional human research ethics policies. Data use is limited to research purposes, on secure computer servers. The principal investigator (VJM) is the contact person for requests to share data.

## Author contributions were as follows

VJM conceived, designed, initiated, and supervised the study; LM led the execution, data collection, analysis, and manuscript preparation. GJB, RRE, JJ, RR, and RP contributed to study design. GJB, NM, JJ, and RP contributed to study execution. All authors critically revised and approved the manuscript.

## Financial support

This work was funded by NIH award K43TW011442 to VJM. LM received financial support through a postgraduate scholarship from the University of Cape Town and the National Research Foundation (NRF South Africa). GJB was supported by postgraduate scholarships from PainSA, the NRF, and the Oppenheimer Memorial Trust. MRH receives funding from the Australian Research Council, the National Health and Medical Research Council, and the Defence Science and Technology Group. RRE is partially supported by NIH Award K24 NS126570.

## Disclosures

GJB receives speakers’ fees for talks on pain and rehabilitation. MRH receives payments from the Department of Education and Department of Science, Industry and Resources; serves as the Chair of the Safeguarding Australia through Biotechnology Response and Engagement Alliance steering committee and the Australian Pain Solutions Research Alliance board; and is a Member of the Prime Minister’s National Science and Technology Council. JAJ has received consultancy fees for research from the University of Maryland and the University of Bern. RP receives payment for lectures on pain and rehabilitation and is an unpaid director of the not-for-profit organisation, Train Pain Academy. VJM also receives payment for lectures on pain and rehabilitation and is an unpaid associate director of the not-for-profit organisation, Train Pain Academy. All other authors declare no conflicts of interest related to this work.

## Notes

### Clinical Trial

NCT04757987

### Clinical Protocols

https://bmjopen.bmj.com/content/12/6/e059723

### Author Declarations

Ethics committee of Faculty of Health Sciences of the University of Cape Town gave ethical approval for this work. Local health authority of the City of Cape Town gave ethical approval for this work

### Summary of Updates

Figure 2 updated in the results section

