## Supplementary files for "Distress is positively associated with induced secondary hyperalgesia in people with suppressed HIV"

Table S1: Bootstrapped covariate-adjusted robust regression models predicting the surface area and magnitude of secondary hyperalgesia (SH), excluding HFS painfulness ratings as a covariate. pid: individual participant code.

|  | **Surface area of SH** | | **Magnitude of SH** | |
| --- | --- | --- | --- | --- |
| *Predictors* | *Estimates* | *CI* | *Estimates* | *CI* |
| Intercept | 8.86 | -23.39 – 41.12 | -2.52 | -12.23 – 7.19 |
| Distress severity | 19.83 ^*^ | 3.40 – 36.25 | 6.36 ^*^ | 1.47 – 11.26 |
| Group (pain-free) | 0.05 | -21.05 – 21.14 | -6.71 ^*^ | -13.33 – -0.08 |
| Baseline mean pinprick ratings | -119.52 | -260.21 – 21.17 | -28.93 | -70.71 – 12.85 |
| Current used for HFS | 1.00 | -1.15 – 3.16 | 0.07 | -0.58 – 0.72 |
| Time difference between distress & SH assessments |  |  | 0.29 ^**^ | 0.09 – 0.48 |
| **Random Effects** | | | | |
| σ^2^ | 178.34 | | 2.99 | |
| τ_00_ | 741.43 _pid_ | | 68.88 _pid_ | |
| ICC | 0.81 | | 0.96 | |
| N | 45 _pid_ | | 45 _pid_ | |
| Observations | 135 | | 135 | |
| Marginal R^2^ / Conditional R^2^ | 0.189 / 0.843 | | 0.246 / 0.969 | |
| ** p<0.05 ** p<0.01 *** p<0.001* | | | | |

Table S2: Summary of conventional models predicting surface area and magnitude of SH. pid: individual participant code.

|  | **Surface area: unadjusted** | | | **Surface area: covariate-adjusted** | | **Magnitude: unadjusted** | | **Magnitude: covariate-adjusted** | |
| --- | --- | --- | --- | --- | --- | --- | --- | --- | --- |
| *Predictors* | *Estimates* | *CI* | | *Estimates* | *CI* | *Estimates* | *CI* | *Estimates* | *CI* |
| Intercept | 4.41 | -20.23 – 29.04 | 10.64 | | -23.70 – 44.97 | -2.01 | -11.50 – 7.48 | -6.09 | -18.06 – 5.87 |
| Distress severity | 16.40 * | 3.14 – 29.66 | 18.06 * | | 2.08 – 34.05 | 4.49 | -0.62 – 9.59 | 7.43 * | 1.77 – 13.08 |
| Group (pain) |  |  | -3.16 | | -24.95 – 18.64 |  |  | -9.35 * | -17.08 – -1.63 |
| Current used for HFS |  |  | -120.25 | | -257.19 – 16.69 |  |  | -24.04 | -71.94 – 23.85 |
| HFS painfulness |  |  | 0.11 | | -0.27 – 0.48 |  |  | 0.09 | -0.06 – 0.24 |
| Time difference between distress & SH assessments |  |  | 0.82 | | -1.28 – 2.92 |  |  | -0.14 | -0.89 – 0.60 |
| Baseline mean pinprick ratings |  |  |  | |  |  |  | 0.28 * | 0.02 – 0.54 |
| **Random Effects** | | | | | | | | | |
| σ^2^ | 242.35 | | | 242.35 | | 43.87 | | 43.87 | |
| τ_00_ | 717.49 _pid_ | | | 715.33 _pid_ | | 103.80 _pid_ | | 81.95 _pid_ | |
| ICC | 0.75 | | | 0.75 | | 0.70 | | 0.65 | |
| N | 45 _pid_ | | | 45 _pid_ | | 45 _pid_ | | 45 _pid_ | |
| Observations | 135 | | | 135 | | 135 | | 135 | |
| Marginal R^2^ / Conditional R^2^ | 0.100 / 0.773 | | | 0.158 / 0.787 | | 0.051 / 0.718 | | 0.241 / 0.735 | |
| ** p<0.05   ** p<0.01   *** p<0.001* | | | | | | | | | |

Table S3: Summary of robust models predicting surface area and magnitude of SH. pid: individual participant code.

|  | **Surface area: unadjusted** | | **Surface area: covariate-adjusted** | | **Magnitude: unadjusted** | | **Magnitude: covariate-adjusted** | |
| --- | --- | --- | --- | --- | --- | --- | --- | --- |
| *Predictors* | *Estimates* | *CI* | *Estimates* | *CI* | *Estimates* | *CI* | *Estimates* | *CI* |
| Intercept | 0.44 | -24.95 – 25.82 | 3.98 | -30.91 – 38.87 | -3.22 | -10.56 – 4.13 | -3.42 | -13.93 – 7.09 |
| Distress severity | 17.62 * | 3.95 – 31.28 | 19.10 * | 2.85 – 35.34 | 4.04 * | 0.09 – 8.00 | 6.24 * | 1.28 – 11.21 |
| Group (pain) |  |  | -1.88 | -24.03 – 20.27 |  |  | -6.92 * | -13.70 – -0.13 |
| Current used for HFS |  |  | -117.83 | -257.02 – 21.36 |  |  | -29.36 | -71.42 – 12.71 |
| HFS painfulness |  |  | 0.13 | -0.25 – 0.51 |  |  | 0.03 | -0.10 – 0.17 |
| Time difference between distress & SH assessments |  |  | 1.07 | -1.06 – 3.20 |  |  | 0.05 | -0.60 – 0.71 |
| Baseline mean pinprick ratings |  |  |  |  |  |  | 0.25 * | 0.02 – 0.48 |
| **Random Effects** | | | | | | | | |
| σ^2^ | 182.59 | | 178.63 | | 2.97 | | 2.99 | |
| τ_00_ | 759.92 _pid_ | | 737.41 _pid_ | | 67.73 _pid_ | | 71.19 _pid_ | |
| ICC | 0.81 | | 0.81 | | 0.96 | | 0.96 | |
| N | 45 _pid_ | | 45 _pid_ | | 45 _pid_ | | 45 _pid_ | |
| Observations | 135 | | 135 | | 135 | | 135 | |
| Marginal R^2^ / Conditional R^2^ | 0.116 / 0.829 | | 0.192 / 0.842 | | 0.084 / 0.962 | | 0.240 / 0.969 | |
| ** p<0.05   ** p<0.01   *** p<0.001* | | | | | | | | |

Table S4: Bootstrapped covariate-adjusted robust models including SH the interaction between distress and group predicting surface area and magnitude of SH

|  | **Surface area of SH** | | **Magnitude of SH** | |
| --- | --- | --- | --- | --- |
| *Predictors* | *Estimates* | *CI* | *Estimates* | *CI* |
| Intercept | 18.73 | -25.44 – 62.89 | 8.12 | -4.37 – 20.61 |
| Distress severity | 8.55 | -16.09 – 33.19 | -1.52 | -8.47 – 5.43 |
| Group (pain) | -31.99 | -91.07 – 27.10 | -29.06 ^***^ | -45.89 – -12.22 |
| Current used for HFS | -111.79 | -248.82 – 25.23 | -21.55 | -60.30 – 17.19 |
| HFS painfulness | 0.14 | -0.24 – 0.51 | 0.01 | -0.12 – 0.13 |
| Time difference between distress & SH assessments | 0.93 | -1.18 – 3.05 | -0.08 | -0.69 – 0.52 |
| Distress x Group (pain) | 17.63 | -14.48 – 49.75 | 13.13 ^**^ | 4.04 – 22.21 |
| Baseline mean pinprick ratings |  |  | 0.31 ^**^ | 0.10 – 0.52 |
| **Random Effects** | | | | |
| σ^2^ | 175.55 | | 3.02 | |
| τ_00_ | 707.95 _pid_ | | 59.86 _pid_ | |
| ICC | 0.80 | | 0.95 | |
| N | 45 _pid_ | | 45 _pid_ | |
| Observations | 135 | | 135 | |
| Marginal R^2^ / Conditional R^2^ | 0.216 / 0.844 | | 0.369 / 0.970 | |
| ** p<0.05   ** p<0.01   *** p<0.001* | | | | |

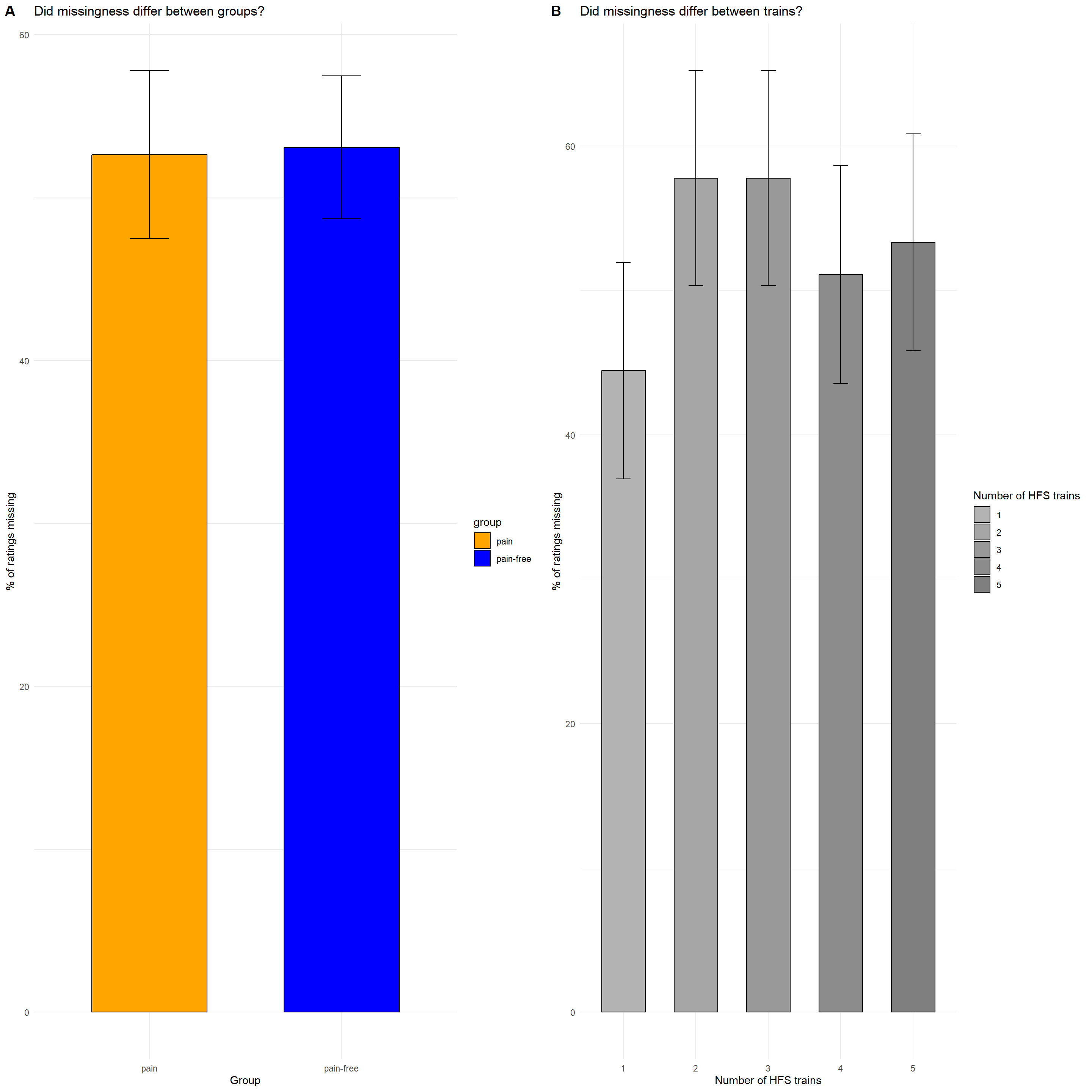

Figure S1: Missingness of HFS ratings did not differ meaningfully by group or train. (A) Proportion of missing ratings (%) in the pain and pain-free groups. (B) Proportion of missing ratings (%) for each HFS train (1–5). Error bars in both panels show standard errors.

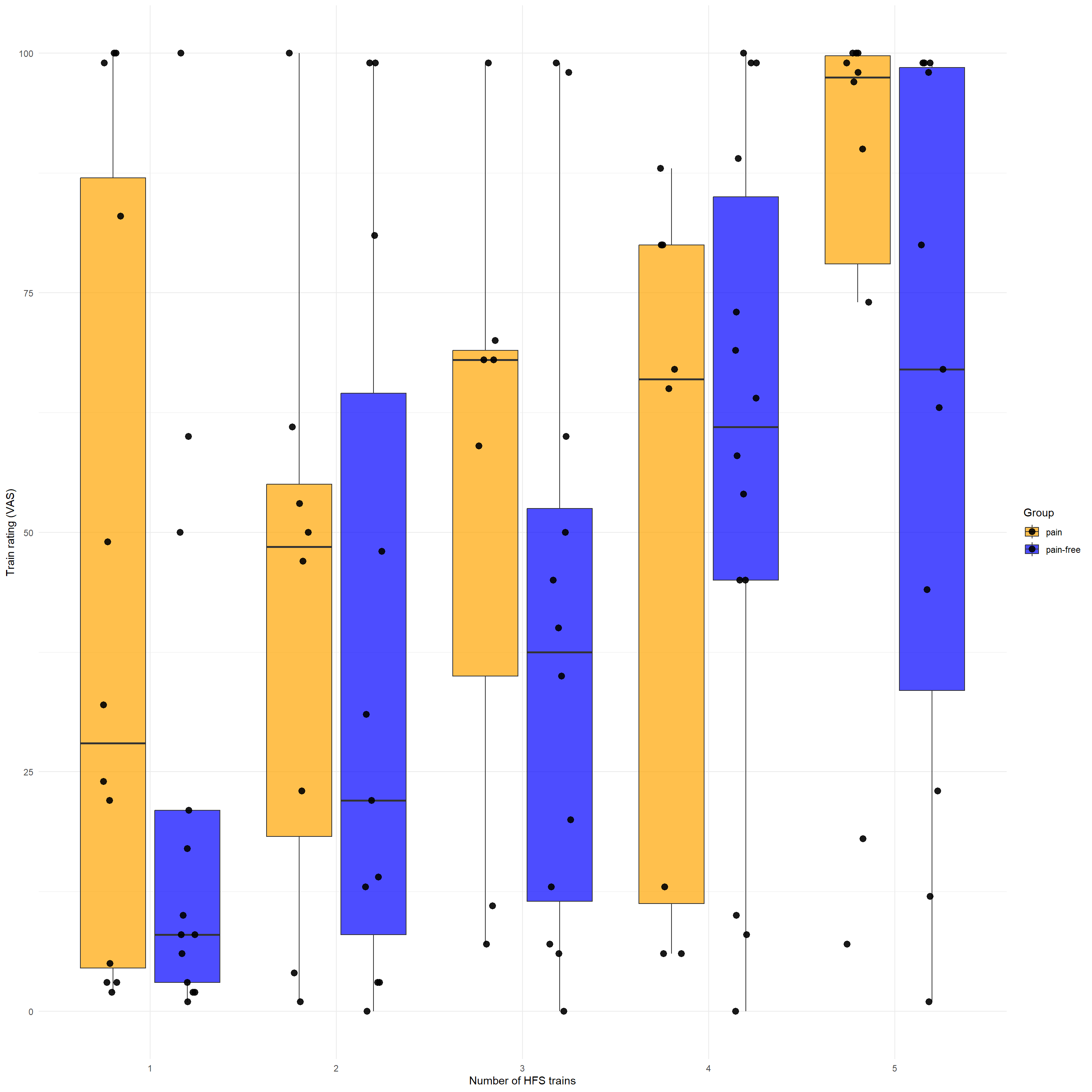

Figure S2: Recorded VAS ratings during each HFS train plotted by group. Black dots represent individual participants’ ratings for each train.

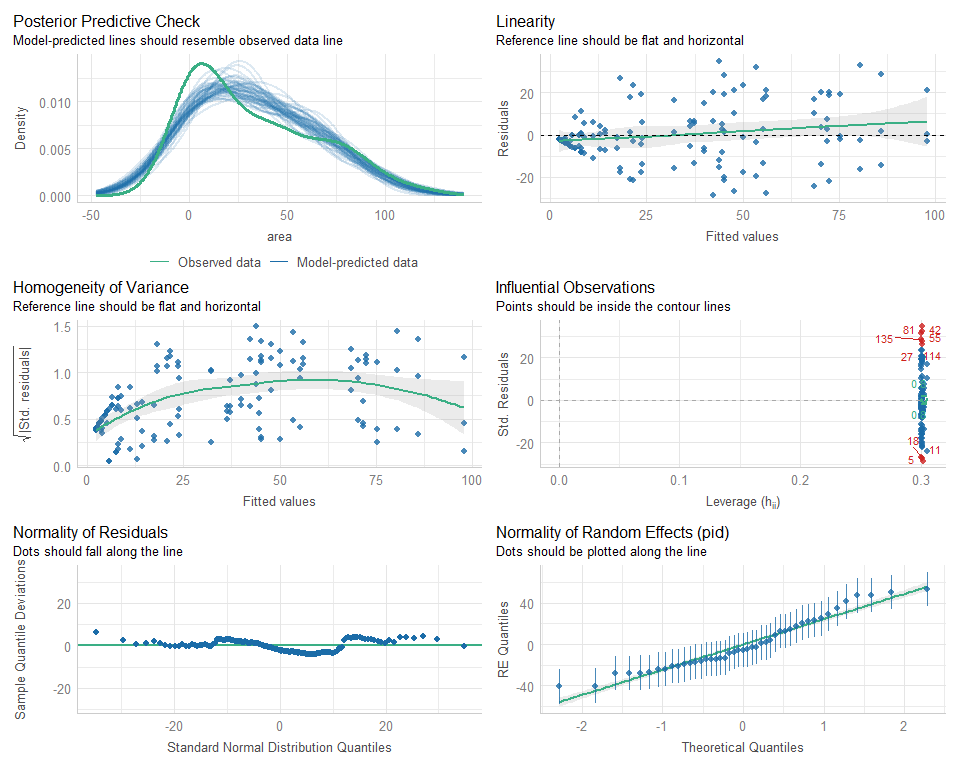

Figure S3: Unadjusted conventional model assumptions for the surface area of SH. The following model assumptions were met; posterior predictive check, linearity, normality of random effects; however, the assumption of influential observations, normality of residuals and homogeneity of variance were not met.

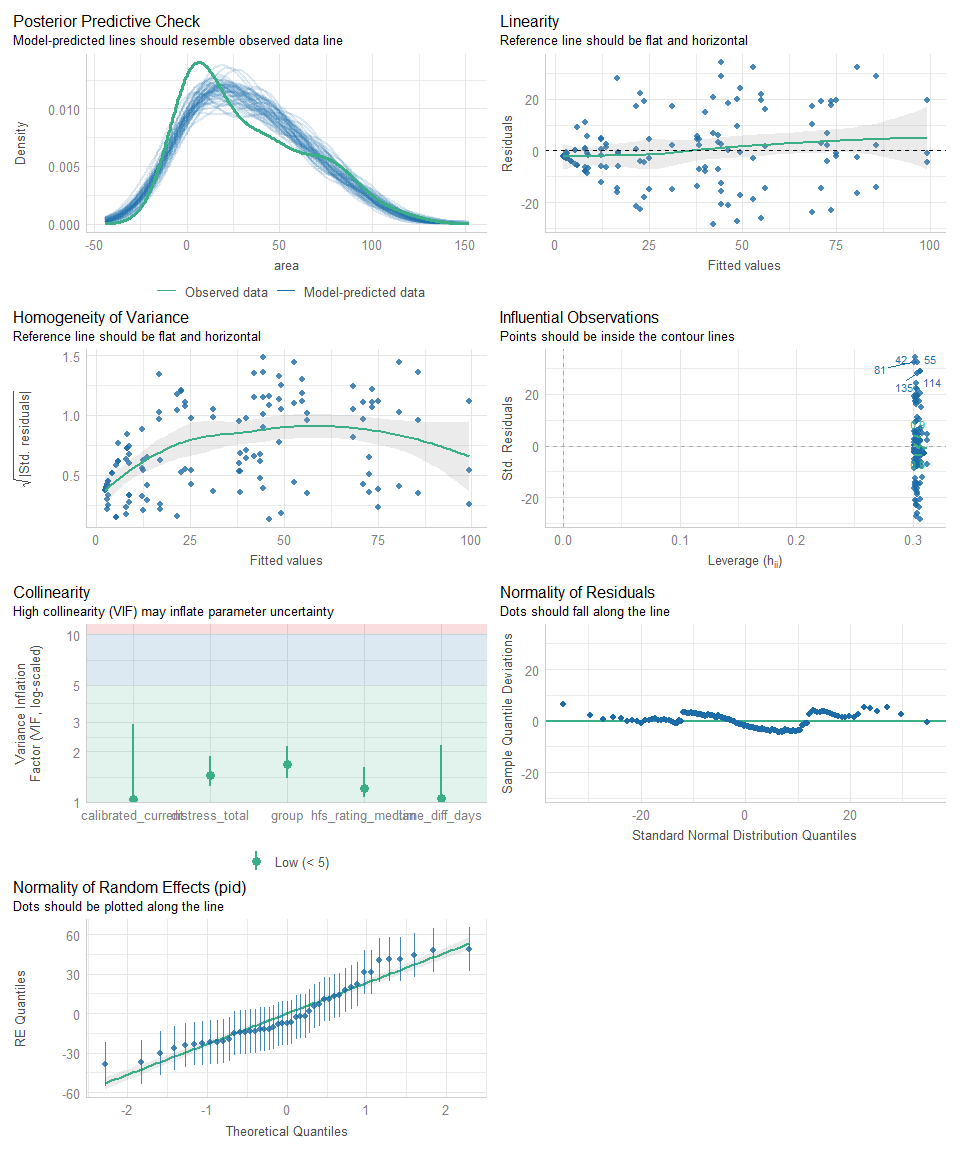

Figure S4: Adjusted conventional model assumptions for the surface area of SH. The following model assumptions were met; posterior predictive check, linearity, collinearity, and normality of random effects; however, the assumption of influential observations, normality of residuals and homogeneity of variance were not met.

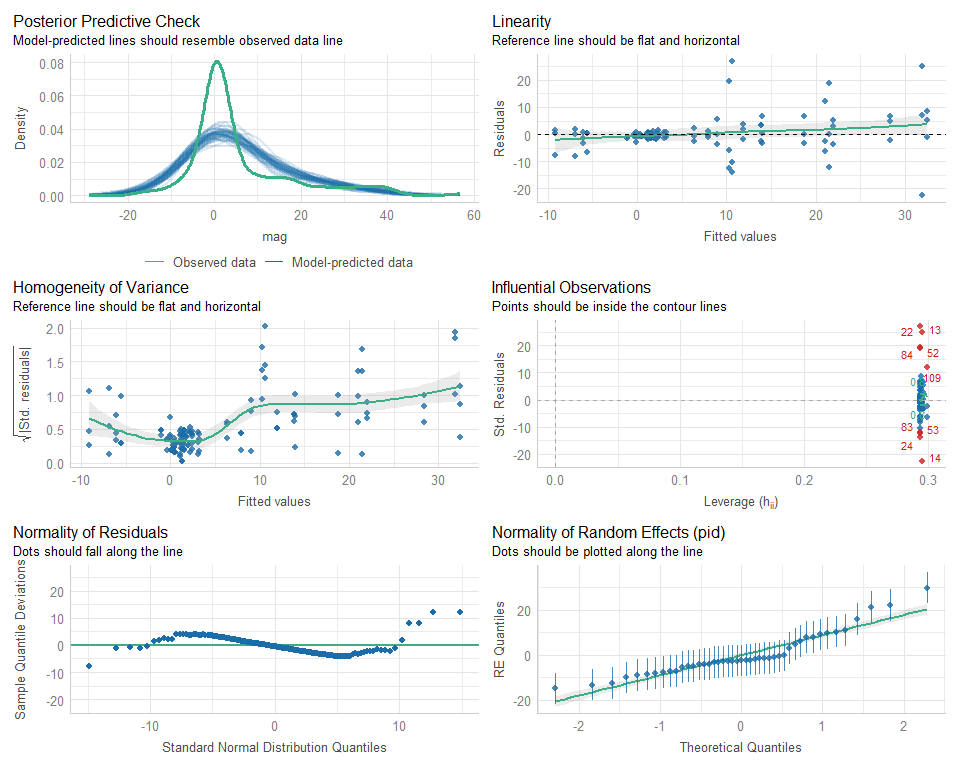

Figure S5: Unadjusted model assumptions for the magnitude of SH. The following model assumptions were met; posterior predictive check, linearity, and normality of random effects; however, the assumption of influential observations, normality of residuals and homogeneity of variance were not met.

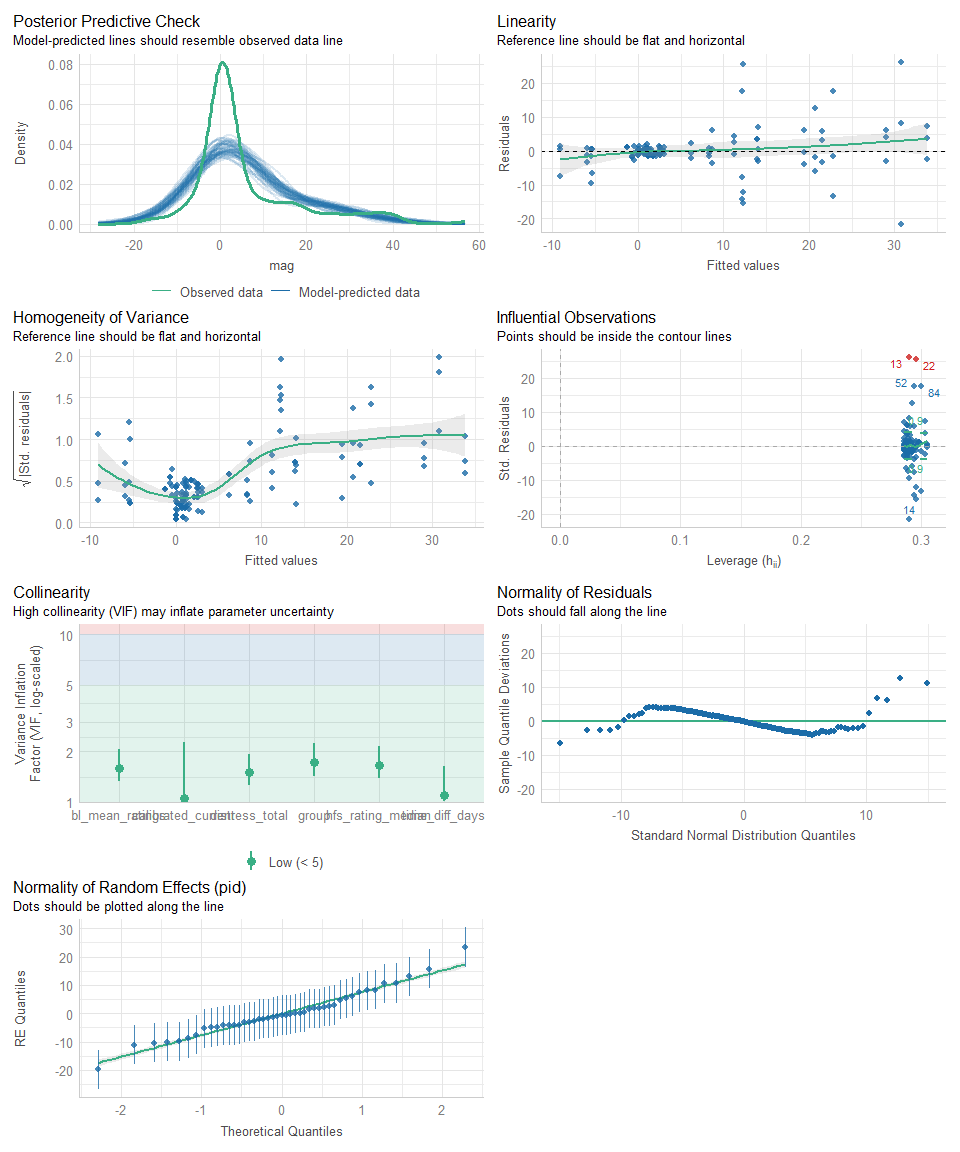

Figure S6: Adjusted conventional model assumptions for the magnitude of SH. The following model assumptions were met; posterior predictive check, linearity, and normality of random effects; however, the assumption of influential observations, normality of residuals and homogeneity of variance were not met.
